# Impact of Access Site on Periprocedural Bleeding, Cerebrovascular, and Coronary Events in High-Bleeding-Risk Percutaneous Coronary Intervention: Findings from the RIVA-PCI Trial

**DOI:** 10.1101/2023.07.19.23292795

**Authors:** Martin Borlich, Uwe Zeymer, Harm Wienbergen, Hans-Peter Hobbach, Alessandro Cuneo, Raffi Bekeredjian, Oliver Ritter, Birgit Hailer, Klaus Hertting, Marcus Hennersdorf, Werner Scholtz, Peter Lanzer, Harald Mudra, Markus Schwefer, Peter-Lothar Schwimmbeck, Christoph Liebetrau, Holger Thiele, Christoph Claas, Thomas Riemer, Ralf Zahn, Leon Iden, Gert Richardt, Ralph Toelg

**Author notes:** **Address for correspondence:** Martin Borlich, MD, Heart Center, Segeberger Kliniken GmbH, Bad Segeberg, Germany, Am Kurpark 1, Bad Segeberg, Schleswig-Holstein, 23795, Germany. **All authors declare:** The article is original, with no portion under simultaneous consideration for publication elsewhere or previously published. All authors have read and approved the submission.

## Abstract

**Background:** The preference for using transradial access (TRA) over transfemoral access (TFA) in patients requiring coronary intervention is based on evidence suggesting that TRA is associated with less bleeding and vascular complications, shorter hospital stays, improved quality of life, and a potential beneficial effect on mortality. We have limited study data comparing both access routes in a patient population with atrial fibrillation undergoing PCI, who have a particular increased risk of bleeding, while AF itself is associated with an increased risk of thromboembolism.

**Methods:** Using data from the RIVA-PCI registry, which includes atrial fibrillation patients undergoing PCI, we analyzed a high-bleeding-risk cohort. These patients were predominantly on oral anticoagulation (OAC) for atrial fibrillation and the PCI was performed via radial or femoral access. Endpoints examined were in-hospital bleeding (BARC 2-5), cerebral events (TIA, hemorrhagic or ischemic stroke) and coronary events (stent thrombosis and myocardial infarction).

**Results:** Out of 1636 patients, 854 (52.2%) underwent transfemoral access (TFA), while 782 (47.8%) received the procedure via transradial access (TRA), including nine patients with brachial artery puncture. Mean age was 75.5 years. Groups were similar in terms of age, sex distribution, atrial fibrillation type, cardiovascular history, risk factors, and comorbidities, except for a higher incidence of previous bypass surgeries, heart failure, hyperlipidemia, and chronic kidney disease (CKD) with GFR<60 ml/min in the TFA group. Clinically relevant differences in antithrombotic therapy and combinations at the time of PCI were absent. However, upon discharge, transradial PCI patients had a higher rate of triple therapy, while dual therapy was preferred after transfemoral procedures. Radial access was more frequently chosen for non-ST-segment elevation myocardial infarction (NSTEMI) and unstable angina pectoris (UAP) cases (NSTEMI 26.6% vs. 17.0%, p<0.05; UAP 21.5% vs. 14.5%, p<0.05), while femoral access was more common for elective PCI (60.3% vs. 44.1%, p<0.05). No differences were observed for ST-segment elevation myocardial infarction (STEMI). Both groups had similar rates of cerebral events (TFA 0.2% vs. TRA 0.3%, p=0.93), but TFA group had a higher incidence of bleeding (BARC 2-5) (4.2% vs. 1.5%, p<0.05), mainly driven by BARC 3 bleeding (1.5% vs. 0.4%, p<0.05). No significant differences were found for stent thrombosis and myocardial infarction (TFA 0.2% vs. TRA 0.3%, p=0.93; TFA 0.4% vs. TRA 0.1%, p=0.36).

**Conclusions:** In high-bleeding-risk (HBR) patients with atrial fibrillation (AF) undergoing PCI for acute or chronic coronary syndrome, utilizing radial access (TRA) resulted in a significant decrease of in-hospital bleeding, while not increasing the risk of embolic or ischemic events compared to femoral access.

## Introduction

Coronary artery disease (CAD) is a major contributor to illness and death worldwide. Patients with CAD and non-valvular atrial fibrillation who are treated with percutaneous coronary intervention (PCI) require oral anticoagulation and additional antiplatelet therapy to prevent thromboembolic events and are therefore exposed to a bleeding risk [1], while atrial fibrillation itself is associated with an increased thromboembolic risk.

According to current guidelines, treatment with novel oral anticoagulants is recommended for these patients, with the optimal additional antithrombotic therapy being the subject of current clinical research. Besides periprocedural anticoagulation and antiplatelet therapy, periprocedural complications can be related to multiple factors. One of the recently studied predictors, especially for in-hospital bleeding events, is the access site utilized for PCI. The preference of TRA over TFA for coronary catheterization has been endorsed by European and American guidelines [2-4]. This is based on evidence suggesting that TRA is linked to a decreased incidence of major bleeding and vascular complications at the access site, shorter hospital stays, and better quality of life [5]. While the majority of data supporting these findings derives from patients undergoing coronary intervention in the setting of acute coronary syndrome (ACS) – including both non-ST-Segment elevation myocardial infarction (NSTEMI) and ST-Segment elevation myocardial infarction (STEMI), data regarding patients with atrial fibrillation undergoing coronary intervention in the setting anticoagulation therapy is scarce [6]. Patients with atrial fibrillation undergoing PCI are more likely to experience adverse outcomes, including in-hospital stroke, compared to those without AF [7]. Moreover, in the recently published SAFARI-STEMI study, there was a trend towards an increased stroke rate in the TRA group compared to the TFA group, with an OR of 2.25 [8].

The objective of this analysis was to examine the rates of in-hospital bleeding (BARC 2-5), cerebral events (TIA, hemorrhagic or ischemic stroke) and coronary events (stent thrombosis and myocardial infarction) with respect to the selected access route for coronary intervention in a large, real-world, prospective cohort of atrial fibrillation patients undergoing PCI (RIVA PCI registry).

## Methods

The “Rivaroxaban in Patients with Atrial Fibrillation Undergoing PCI” (RIVA-PCI) was a prospective, non-interventional, multicenter observational study of consecutive patients with non-valvular AF undergoing PCI that examined the current antithrombotic treatment regimen in Germany after PCI in a 14-month follow-up of a real-world population registry. This cohort was not limited to taking specific OACs or platelet aggregation inhibitors. Patients were enrolled between 2018 and 2020 and data have been published recently [9]. The inclusion criteria were: signed written informed consent before or after PCI; age ≥18 years; knows or newly diagnosed non-valvular AF; PCI with stent implantation during index hospital stay. The exclusion criteria were: participation in any randomized trial influencing the antithrombotic therapy, and patients who were compulsorily detained for treatment of either a psychiatric or physical illness. Informed consent could be signed either before or after PCI. In this sub-analysis, patients from the aforementioned study were compared with respect to the access site utilized for vascular access during the index PCI.

Endpoints examined were in-hospital bleeding (BARC 2-5), cerebral events (TIA, hemorrhagic or ischemic stroke) and coronary events (stent thrombosis and myocardial infarction).

Stroke was characterized as a sudden onset of neurological dysfunction, either focal or global, due to vascular damage of blood vessels in the brain, spinal cord, or retina caused by either hemorrhage or infarction. Ischemic stroke was defined as sudden onset of focal dysfunction of the central nervous system tissue, be it cerebral, spinal, or retinal, caused by infarction. Hemorrhagic stroke refers to a sudden onset of cerebral or spinal dysfunction, either focal or global, caused by intraparenchymal, intraventricular, or subarachnoid hemorrhage. In cases where there was insufficient information to categorize a stroke as either ischemic or hemorrhagic, it was defined as an undetermined stroke. Bleeding episodes were classified according to the Bleeding Academic Research Consortium (BARC) Bleeding Definitions in this analysis[9].

The study was approved by the ethics committee of Landesärztekammer Rheinland-Pfalz, Germany (no. 837.448.17).

## Statistical analysis

The data for this study were collected and analyzed from the RIVA-PCI study database. The incidence of in-hospital bleeding and cerebral events was compared between the femoral and radial access groups using chi-squared tests or Fisher’s exact tests, as appropriate. Patient characteristics and baseline variables were compared between both groups using t-tests or Mann-Whitney resp. Wilcoxon test for continuous variables and chi-squared tests for categorical variables. A p-value <0.05 was considered statistically significant. Binary, categorical, and ordinal parameters are described using absolute numbers and percentages, while continuous/numerical variables are described by means of standard statistics. The sample size for this sub-analysis was determined based on the overall sample size of the RIVA-PCI study, which included a total of 1636 patients. Statistical calculations were performed using IBM SPSS Statistics and GraphPad Prism software.

## Results

## Baseline characteristics

In the RIVA-PCI study, a total of 1636 patients with atrial fibrillation undergoing percutaneous coronary intervention (PCI) were enrolled, of which 854 (52.2%) received a transfemoral access (TFA), while 782 (47.8%) underwent the procedure via the transradial access (TRA) including nine patients with a brachial artery puncture. These 782 patients were summarized as radial access group. The mean age of patients in the femoral group was 75.1 years, while the mean age in the radial access group was 75.9 years.

Our analysis revealed no significant differences regarding baseline characteristics between both groups, except for a higher prevalence of heart failure (48.1% vs. 38.7%, p<0.05), a history of previous bypass surgery (15.6% vs. 7.7%, p<0.05), hyperlipidemia (64.5% vs. 58.2%, p<0.05), GFR<60 ml/min (49.4% vs. 42.1%, p<0.05) and patients with dual therapy at baseline (16.3% vs. 12.3%, p<0.05) in the femoral access group.

When considering the antithrombotic therapy of these patients with atrial fibrillation undergoing PCI, there was no difference between the TRA and TFA groups regarding those who did not receive any antithrombotic therapy prior to treatment, single antiplatelet agent, novel oral anticoagulants (NOAC) only, or Vitamin K antagonist (VKA) only. However, a significant number of patients in the TFA group had dual a therapy with acetylsalicylic acid (ASA)P2Y12-inhibitor + VKA/NOAC (16.3% vs. 12.3%, p<0.05) (**Table 1**).

**Table 1.**
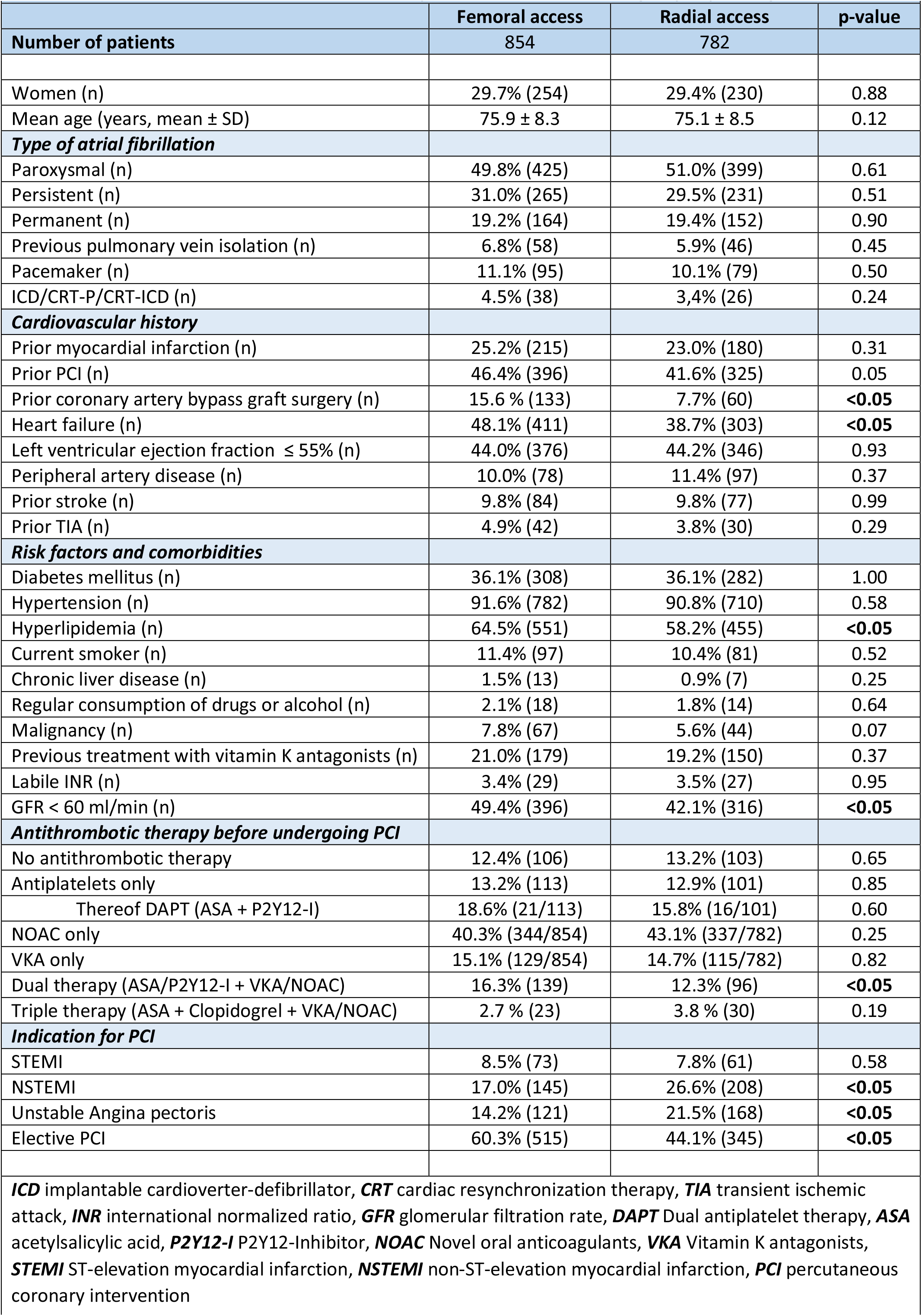
Baseline characteristics of the 1636 patients enrolled in the registry depending on access route.

In cases of non-ST-segment elevation myocardial infarction (NSTEMI) and unstable angina pectoris (UAP), the radial approach was more frequently chosen (NSTEMI 26.6% vs. 17.0%, p<0.05; UAP 21.5% vs. 14.5%, p<0.05), while the femoral approach was more commonly used for elective PCI (60.3% vs. 44.1%, p<0.05). There was no difference between the two groups in cases of ST-segment elevation myocardial infarction (STEMI) (**Table 1**). A vascular closure device was used more frequently after femoral puncture (73.8 % vs. 17.4%, p<0.05).

In both groups, the median CHA_2_DS_2_-VASc score was 5, while the median HAS-BLED score was 3 in the femoral access group and 2 in the radial access group. **Figure 1** provides a detailed overview of the distributions of both scores.

**Figure 1:**
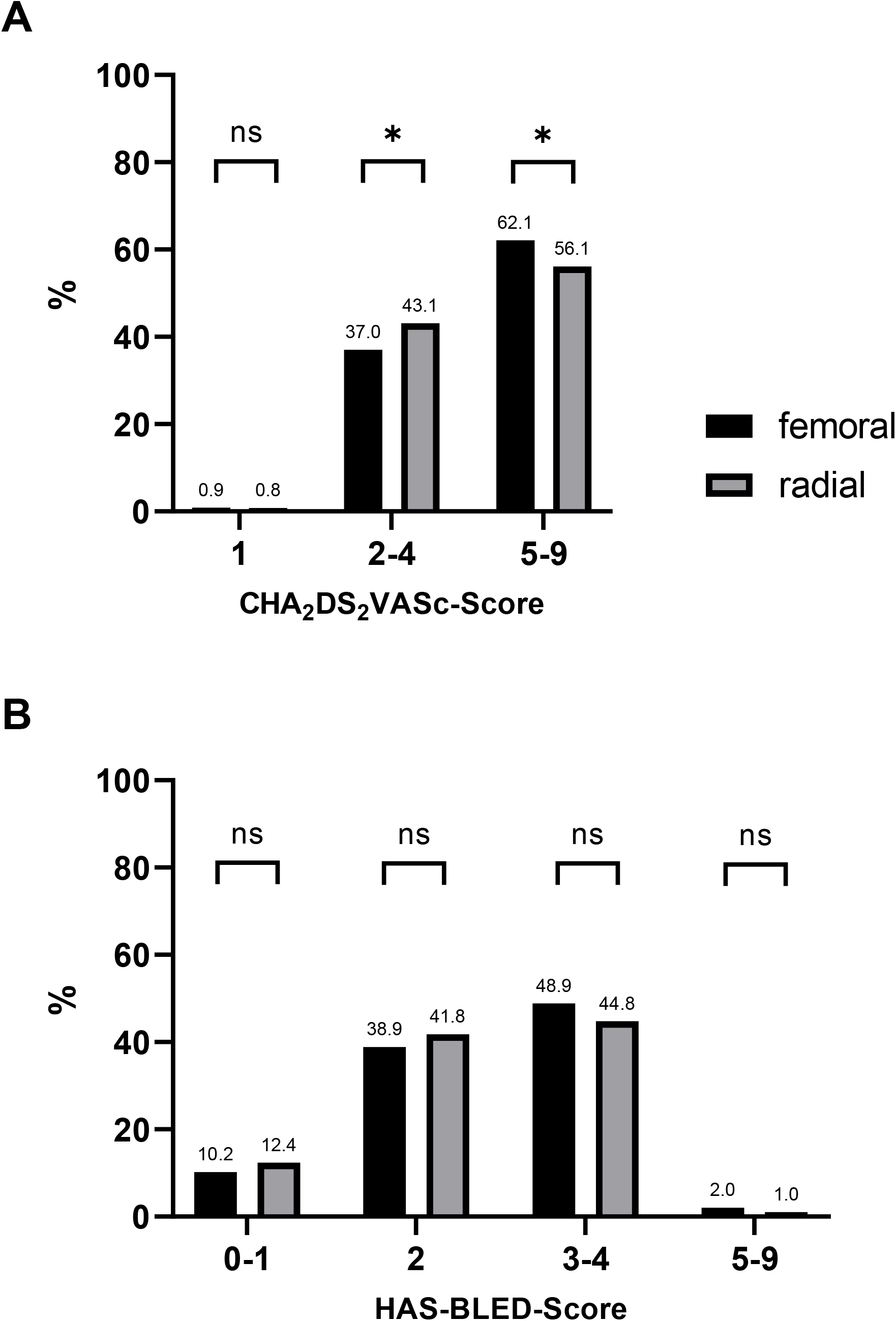
Distribution of CHA2DS2-VASc-Score (A) and HAS-BLED Score (B); * = statistically significant difference (p<0.05), ns= not significant

### Examined endpoints

Our results show that there was no significant difference in the incidence of in-hospital stroke or TIA between the two groups (two ischemic strokes in the TFA group and one ischemic stroke and one stroke of unknown type in the TRA group). There was a significant difference in the incidence of bleeding events, with significantly more bleeding that requires transfusion or surgical therapy (BARC 3) in the TFA group compared to the TRA group (1.5% vs. 0.4%; p<0.05). Fatal bleeding events did not occur (**Table 2**). There were no differences observed between the femoral access and radial access groups in terms of coronary events. The incidence of myocardial infarction was 0.4% in the femoral access group and 0.1% in the radial access group (p=0.36), and the occurrence of stent thrombosis was 0.2% in the femoral access group and 0.3% in the radial access group (p=0.93). Additionally, the rate of all-cause mortality was similar between the two groups (0.9% [TFA] vs. 0.5% [TRA], p=0.31).

**Table 2.** In-hospital bleeding, cerebral and coronary events until discharge depending on access route for PCI.

|  |  |  |  |
| --- | --- | --- | --- |
|  | **Femoral access** | **Radial access** | **p-value** |
| **Number of patients** | 854 | 782 |  |
| Days between PCI and discharge, if alive (mean ± SD) | 4.4 ± 5.6, N=846 | 4.0 ± 4.8, N=778 | 0.12 |
| *TIA/Stroke* |  |  |  |
| Ischemic | 0.2 % (2/854) | 0.1 % (1/782) | 0.62 |
| Hemorrhagic | 0.0 % (0/854) | 0.0 % (0/782) | --- |
| Unknown type | 0.0 % (0/854) | 0.1 % (1/782) | 0.30 |
| TIA | 0.0 % (0/854) | 0.0 % (0/782) | --- |
| **Overall (stroke & TIA)** | **0.2 % (2/854)** | **0.3 % (2/782)** | **0.93** |
| *Bleeding* |  |  |  |
| **BARC 5 (Fatal)** | 0.0 % (0/854) | 0.0 % (0/782) | --- |
| BARC 4 (CABG related) | 0.0 % (0/854) | 0.0 % (0/854) | --- |
| BARC 3 (Surgery or transfusion) | 1.5 % (13/854) | 0.4 % (3/782) | **< 0.05** |
| BARC 3c (Intracranial, -spinal, -ocular) | 0.0 % (0/854) | 0.1 % (1/782) | 0.30 |
| BARC 3b (Overt, Hb drop ≥5g/dl) | 0.5 % (4/854) | 0.1 % (1/782) | 0.21 |
| BARC 3a (Overt, Hb drop <5g/dl) | 1.1 % (9/854) | 0.1 % (1/782) | **< 0.05** |
| BARC 2 (Nonsurgical treatment) | 1.2 % (10/854) | 0.8 % (6/782) | 0.41 |
| **Overall (BARC 2-5)** | **4.2 % (36/854)** | **1.5 % (12/782)** | **< 0.05** |
| *Coronary events* |  |  |  |
| Stent thrombosis | 0.2 % (2/854) | 0.3 % (2/782) | 0.93 |
| Myocardial infarction | 0.4 % (3/854) | 0.1 % (1/782) | 0.36 |
| **Overall (Stent thrombosis & Myocardial infarction)** | **0.6 % (5/854)** | **0.4 % (12/782)** | **0.73** |

The average length of the hospital stay until discharge for patients undergoing PCI was four days in both the femoral access and radial access group. Upon discharge, transradial PCI patients had a higher rate of triple therapy (33.2% vs. 19.3%, p<0.05), while dual therapy was preferred after transfemoral procedures (72.7% vs 59.2%, p<0.05).

## Discussion

This sub-analysis of the RIVA-PCI trial [9] aimed to investigate in-hospital bleeding, cerebral and coronary event rates depending on femoral or radial access for coronary intervention in patients with atrial fibrillation. Meta-analyses showed that in patients undergoing coronary angiography and PCI, radial access (TRA) is associated with a significant reduction in the risk of bleeding, vascular complications, and mortality compared to femoral access (TFA). The risk of stroke or MI was comparable in patients with radial or femoral access [6, 10]. A considerable number of these studies investigated coronary interventions performed during an acute coronary syndrome (ACS). Moreover, it is still controversial whether the benefits of radial versus femoral access for coronary angiography and percutaneous coronary interventions (PCI) are based on the choice of access site itself, the experience of the operator, or other mechanisms.

The data underlying the present investigation derives from the RIVA PCI registry and therefore addresses a cohort of patients with atrial fibrillation who underwent PCI, both in acute and chronic coronary syndrome. The study was conducted at multiple centers allowing any significant differences in the selected parameters of bleeding and cerebral event rate to be attributed with high confidence to the selected access route. The study focused on a homogeneous patient population with atrial fibrillation, which adds to the growing evidence comparing these two access sites in patients undergoing PCI.

In this study, the use of femoral access for PCI was associated with a higher bleeding rate (BARC 2-5) compared to the radial access group (4.2 % vs. 1.5 %, p<0.05). This is caused mainly by significantly more bleeding that requires transfusion or surgical therapy (BARC 3). These differences were observed peri-interventionally. This is particularly remarkable considering that in the TRA group, significantly more patients were prescribed triple therapy at the time of discharge compared to the TFA group, which predominantly received dual therapy. Our results are consistent with the data from a recently published meta-analysis of 31 randomized controlled trials (RCTs) comparing radial versus femoral access sites for coronary angiography and PCI. In that meta-analysis, radial access was associated with a significant reduction in major bleeding compared to femoral access (OR 0.53, 95%CI 0.42–0.66). These findings were consistent regardless of clinical characteristics or whether coronary angiography was performed with or without PCI [6]. A similar OR of 0.55 was calculated in the meta-analysis by Gargiolu et al., which was published in 2022 [5]. The odds ratio (OR) for the defined endpoint of BARC 2-5 bleeding in our analysis is 0.36 [95% CI 0.18-0.68]. These findings should be taken into consideration when selecting the optimal access site for PCI, particularly in patients who may be at higher risk for bleeding complications. In our analysis, the number needed to treat (NNT) is 37 to avoid a relevant bleeding that needs to be treated (BARC 2-5). Regardless of the puncture technique used and any additional measures taken to limit vascular complications, there is mounting evidence that the superior safety profile of TRA over TFA persists. While advancements in ultrasound guidance and micropuncture needles may decrease the incidence of vascular complications after TFA, TRA still appears to yield better outcomes in terms of safety [11, 12].

The incidence of TIA and stroke, however, was similar between both groups. Although the TRA is associated with an increased rate of subclinical cerebral emboli, no difference in the rates of clinically relevant strokes has been demonstrated in large randomized studies [8, 13, 14]. This may be due to the low incidence of periprocedural strokes. With radial access, there could be an increased risk of stroke due to catheter manipulation in the extracranial cervical vessels, but on the other hand, this risk also exists with femoral access and catheter manipulation in the aortic arch, especially in the presence of plaques in the thoracic aorta. In our study, two strokes occurred in both groups, two ischemic strokes in the femoral access group (0.2%) and one ischemic stroke and one stroke of unknown type in the patient cohort with radial access (0.3%). As no systematic neurological examination was performed post-intervention, as in most of the other studies, the true event rate may be higher. The following consideration should be mentioned at this point: even to investigate a 50% increase in stroke rate with a stroke incidence of 0.2% and a statistical power of 80%, nearly 80,000 patients would be required.

The findings of this study also suggest that the selection of access site, either transfemoral (TFA) or transradial (TRA), did not significantly impact the risk of stent thrombosis or myocardial infarction. The occurrence of stent thrombosis was observed in 0.2% of patients in the TFA group and 0.3% of patients in the TRA group, with no statistically significant difference between the two groups (p=0.93). Similarly, the incidence of myocardial infarction was 3 cases in the TFA group and 1 case in the TRA group, with no significant difference between them (p=0.36). This is consistent with previous research indicating comparable outcomes between the femoral and radial approaches in terms of coronary events [5]. The studied data also revealed that there was no difference in all-cause mortality between both groups. Until discharge, 12 out of 1636 patients died, eight in the TFA group (six due to cardiac death, 2 unknown) and four in the TRA group (three due to cardiac death, 1 unknown). There was no statistically significant difference between both groups (0.9% vs. 0.5%, p=0.31). The mean length of stay after PCI until discharge was four days, and there was no significant difference between both groups. A meta-analysis published in 2022 showed that all-cause mortality was lower with TRA compared to TFA at 30 days (1.6% versus 2.1%; HR, 0.77 [95% CI, 0.63-0.95]; p=0.012) with a number needed to treat to benefit (NNTB) of 214. Landmark analyses demonstrated that the benefit in favor of TRA was mainly observed in the first few days after the index intervention (0.5% versus 0.8%; HR 0.64 [95% CI, 0.46-0.90]; 0.010)[5]. Our data roughly correspond to these results, although our sample size was significantly smaller, thereby increasing the susceptibility to a type II error. Major bleeding can be considered as a mediator of all-cause mortality (indirect effect), especially in patients with moderate or severe baseline anemia [15].

The access route was chosen by the interventionalist and therefore it is possible that sicker, possibly more complex patients were more likely to receive their PCI through the femoral access. As shown in Table 1 and Figure 2, patients with high CHA_2_DS_2_VASc-score (5-9) were more likely to receive a femoral approach. Patients treated via the TFA also had a higher prevalence of heart failure and previous CABG surgeries. Interventionalists may have opted for the femoral access route more frequently for sicker patients.

**Figure 2:**
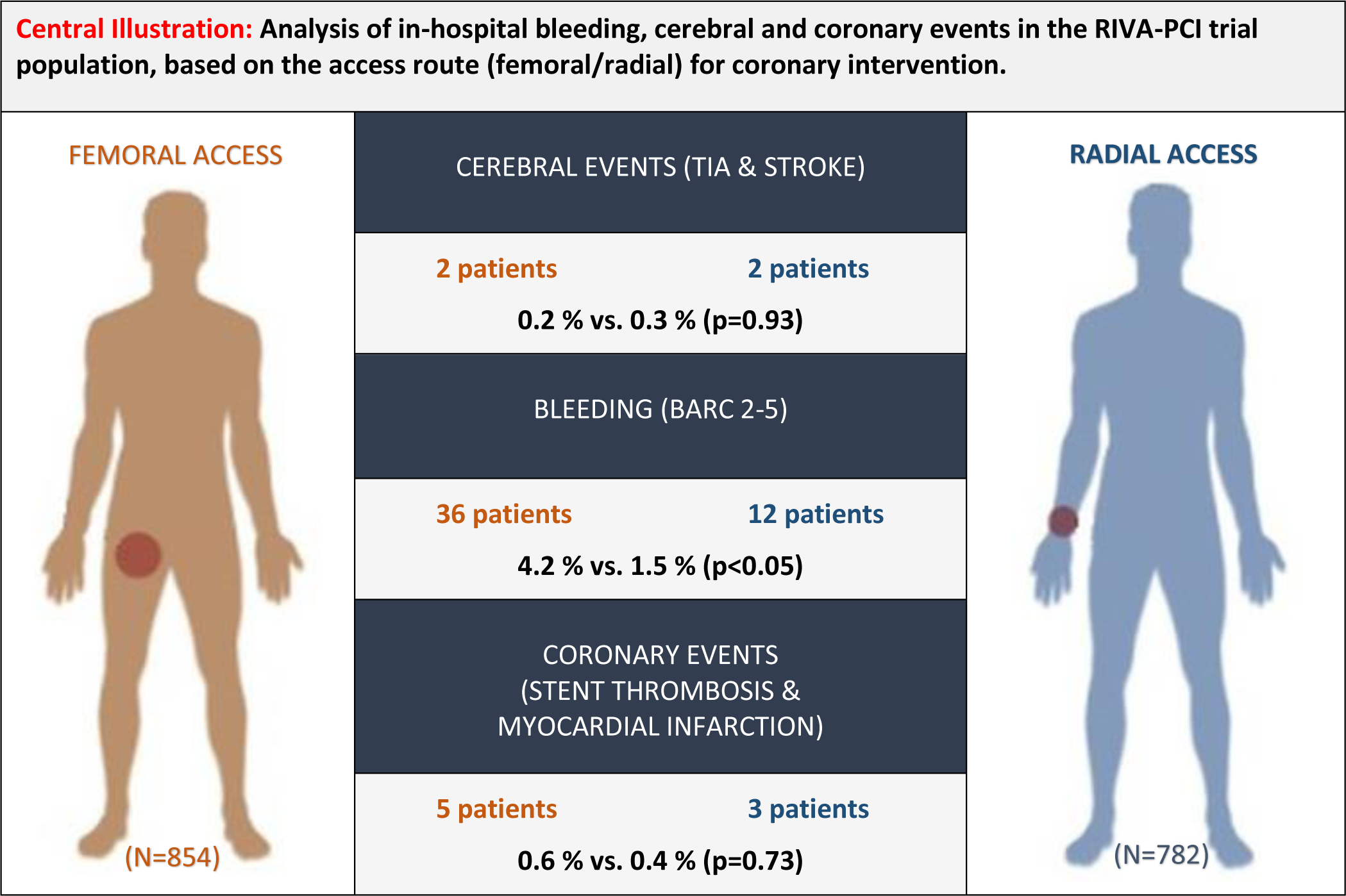
Analysis of in-hospital bleeding, cerebral and coronary events in the RIVA-PCI trial population, based on the access route (femoral/radial) for coronary intervention. In addition to the absolute numbers of affected patients, event rates are given as percentages. The case number of patients with femoral access (TFA) was 854, that with radial access (TRA) 782.

Overall, this study suggests that using femoral access during PCI in patients with atrial fibrillation may be associated with a higher in-hospital bleeding rate compared to radial access, but there is no difference in cerebral or coronary event rate. These data is in line with previously published results of comparisons between both access routes, which, however, predominantly originated from patients who received PCI for acute coronary syndrome and were not all affected by atrial fibrillation.

Therefore, further research is needed to confirm these results and to determine which patients are most likely to benefit from each access site.

### Study limitations

The RIVA PCI study was a registry-based observational study. Registry studies have some limitations that should be considered to interpret the results appropriately. These limitations include selection bias, where certain groups of patients may be less likely to be included in the registry or may be less willing to participate. Additionally, information bias is possible, although this registry had high accuracy and completeness of data. Like all registry studies, it is not easy to account for the influence of other factors on the outcomes being studied (confounding). The calculations performed in this study were retrospective, provide valuable information, and generate hypotheses, but randomization into TRA and TFA groups as in an RCT was not performed. Furthermore, the group sizes were not a priori powered to detect a small difference in mortality, coronary or cerebral events.

### Conclusions

In conclusion, this sub-analysis of the RIVA-PCI trial demonstrates that using radial access for coronary intervention in patients with atrial fibrillation is associated with a significant reduction in the incidence of in-hospital bleeding (BARC 2-5) compared to femoral access. Additionally, there was no significant difference in the occurrence of cerebral events (TIA, hemorrhagic or ischemic stroke) or coronary events (stent thrombosis or myocardial infarction) between the two groups (**Figure 2**). Notably, this study included patients with both acute and chronic coronary syndromes undergoing PCI. These findings support the use of transradial access in patients with atrial fibrillation undergoing PCI, particularly in those at higher risk for bleeding complications.

## Data Availability

The data used in this study are available upon request. However, due to privacy and ethical considerations, access to the data is subject to approval from the appropriate regulatory bodies and the RIVA-PCI Trial investigators.

## Disclosures

None.

## Notes

### Competing Interest Statement

The authors have declared no competing interest.

### Funding Statement

This research received no specific funding from any external sources.

### Author Declarations

The ethics committee of Landesaerztekammer Rheinland-Pfalz (Germany) gave ethical approval for this work (no. 837.448.17).

